# Optimizing diagnostic strategy for novel coronavirus pneumonia, a multi-center study in Eastern China

**DOI:** 10.1101/2020.02.13.20022673

**Authors:** Jing-Wen Ai, Hao-Cheng Zhang, Teng Xu, Jing Wu, Mengqi Zhu, Yi-Qi Yu, Han-Yue Zhang, Yang Li, Xian Zhou, Zhongliang Shen, Guo-Qing Zang, Jie Xu, Wen-Jing Chen, Yong-Jun Li, De-Sheng Xie, Ming-Zhe Zhou, Jing-Ying Sun, Jia-Zhen Chen, Wen-Hong Zhang

## Abstract

COVID-19 caused by a novel coronavirus SARS-CoV-2 emerged in Wuhan, Hubei province since December 2019, and caused a rapid outbreak throughout China and globally. Cities outside Hubei are also facing great challenge and require implementing of effective and feasible strategy in precision diagnosing novel coronavirus pneumonia (NCP).

We described a multicenter prospective study on diagnostic strategy of suspected NCP patients from January 22^nd^ to February 9^th^, 2020 in Eastern China cities. Nasopharyngeal swabs were collected from the patients. The epidemiological characteristics, clinical symptoms, laboratory assessments, and computed tomographic (CT) scans were obtained. Pathogen screen were performed including RT-PCR, multiplex PCR, rapid flu antigen tests and mNGS.

We enrolled 53 suspected NCP patients, among whom 20 were laboratory-confirmed. Fourteen (70%) and 3 (15%) patients were positive for the first and second SARS-CoV-2 RT-PCR test, respectively. All NCP patients were positive for mNGS. Chest CT images and the symptoms of early stage NCP patients were similar to other viral pneumonia patients. We identified 11 of 20 co-infections in NCP cases, including regular respiratory virus, fungi and bacteria synchronously. Genomic analysis showed that 8 of 10 cases had no mutation in virus genome, while 2 cases had only one single mutation in N gene.

Our study discovered that a combination of chest CT, SARS-CoV-2 RT-PCR and multi-plex PCR is recommended in regions outside Hubei province. Co-infection of other pathogens with SARS-CoV-2 exists and should be acknowledged. Repeated sampling, change of specimen type or metagenomics sequencing could further facilitate during critical clinical cases.

## Introduction

Since December 2019, clusters of unexplained pneumonia emerged in Wuhan, China, and rapidly spread throughout China and globally. In early January, Chinese health authority announced the causative agent was isolated to be a novel coronavirus, and the whole genome of which was published^1-3^. The virus was initially named by WHO as the 2019 novel coronavirus and was renamed in 11^th^ Feb 2020 as SARS-CoV-2^4^. Until February 13^th^, 2020, the total reported confirmed corona virus disease 2019 (COVID-19) cases have reached 48,296 within China, and 490 cases in other 24 countries globally^5,6^, and the increasing number of cases and widening geographical spread have raised concerns internationally^7^.

Apart from the transmission pattern and characteristics of this virus, one of the reasons for the initial rapid growth of infected-patient numbers was insufficient ability to rapidly detect such a huge amount of suspected cases, especially in a city with more than 10 million populations. This could then led to the untimely discover and quarantine patients infected with SARS-COV-2, further aggravate the epidemics. Outside Hubei Province, sporadic and clustered cases have continuously reported within China, and most of the provinces and municipalities had confirmed around 50-1000 cases. Other countries including USA, South Korea, Japan and etc, also had sporadic and clustered reports^5,6,8-10^. Currently, the epidemics control outside Hubei province and other countries has entered into a critical stage. To control and eliminate the epidemics is still the main task till now; thus to prevent a single spark to start a prairie fire, as what we had observed in Wuhan, is critical. Therefore, optimizing diagnostic strategy is urgent and essential for the future global SARS-COV-2 control and prevention.

Chest computerized tomography (CT) is an important method to identify lung lesions. A study involving 1099 patients have shown that around 76.4% of the laboratory-confirmed SARS-COV-2 patients had novel coronavirus pneumonia (NCP) on admission, and usually exhibited typical radiological finding of the ground-glass opacity (50%) or bilateral patchy shadowing (46%)^11^. However, the specificity of chest CT is relatively low. Currently, the golden standard for the NCP diagnosis is a positive result in real-time reverse-transcriptase polymerase-chain-reaction (RT-PCR) assay or metagenomics sequencing for respiratory specimens, including nasal and pharyngeal swab specimens, sputum, and bronchoalveolar lavage fluid^12^. Importantly, RT-PCR is required to be performed at least twice for suspected cases on both admission day 0 and day 2 to achieve two negative results in order for the patients to be released. However, recently, physicians around china has reported cases of NCP that had 2 or even 3 continuous negative RT-PCR results for nasopharyngeal and throat swab specimens before finally laboratory-confirmed^13^. According to the recent “Diagnosis and Treatment Guideline for New Coronavirus Pneumonia (the fifth edition), China”, CT imaging results were used as the clinical diagnostic criteria for NCP, but strictly limited in Hubei Province^14^. This phenomenon raised national concerns and discussions about how to improve the diagnostic strategy of novel coronavirus pneumonia (NCP) under the current circumstances.

Considering the high incidence of other viral lung diseases and seasonal cold in winter, cities outside Hubei province also face a great challenge in implementing effective and feasible strategy in precision diagnosing NCP patients. Missed diagnosed NCP patients lead to exposure risks to communities and health care workers, while over-diagnosed of suspected patients means occupation of medical resources and increasing cross-infection of these pneumonia patients (usually arranged in the same ward with NCP patients), leading to unnecessary panic and gradual depletion of medical resources. This article is a pilot study and aims to detail describe the pathogens of suspected NCP patients in the eastern-China region, to compare the efficacy of different diagnostic approaches in NCP diagnosis, and to provide evidence for future strategic diagnosis in regions outside Hubei Province.

## Methods

### Patients

We described a multicenter prospective study on the diagnosis of suspected SARS-COV-2 pneumonia (NCP) from January 22^nd^ to February 9^th^, 2020 in Eastern China cities. Cases were diagnosed based on the WHO interim guidance^15^. According to the Chinese guideline, suspected NCP cases were identified as having pneumonia after chest CT (with one of the two following criteria met: fever or respiratory symptoms, normal of decreased white blood cell counts/ decreased lymphocytes counts), and a travel history or contact with patients with fever or respiratory symptoms from Hubei Province or confirmed cases within 2 weeks. A confirmed case with NCP was defined as a positive SARS-COV-2 nucleotides result either by metagenomic sequencing or RT-PCR assay for nasopharyngeal swab specimens^14^. Severe cases are defined as any one of the followings: 1) Respiratory distress, respiratory rates ≥ 30 per minute; 2) Pulse oxygen saturation ≤93% on room air 3) Oxygenation Index (PaO2/FiO2) ≤300mmHg. Oral consents were obtained from all the patients and the study was approved by the ethics review committee of Huashan Hospital affiliated to Fudan University.

### Data collection

The epidemiological characteristics (including recent exposure history), clinical symptoms and signs, laboratory assessments and CT scans were obtained with data collection forms from electronic medical records. Laboratory assessments consisted of complete blood count, liver function, C-reactive protein and direct antigen Flu A+B test.

### Sample collection and laboratory test

Nasopharyngeal swab samples were collected for extracting total RNA from all included suspected patients on the admission day to fever clinic (DAY 0). After collection, the nasopharyngeal swabs were placed into a collection tube with 3 ml of virus transport solution (Copan UTM^®^: Viral Transport, USA). One-milliliter each of the sample were sent for SARS-COV-2 RT-PCR to both key laboratory of pathogenic microorganisms of Huashan Hospital and Jingan District Center for Disease Prevention and Control^11^. If the first RT-PCR result was negative, the second nasopharyngeal sample of observing patients would be collected on DAY 3 for RT-PCR test again.

At the meantime, 300μl of UTM were sent for a fast, simultaneous, multiplexed real-time reverse transcription nucleic acid amplification assay for qualitative detection and identification of multiple respiratory viral and bacterial nucleic acids in nasopharyngeal swabs using Qiagen ResPlex II V2.0 kit, which including more than 22 pathogens: Adenovirus, Coronavirus 229E, Coronavirus HKU1, Coronavirus NL63, Coronavirus OC43, Human Metapneumovirus A+B, Influenza A, Influenza A H1, Influenza A H3, Influenza A H1N1/pdm09, Influenza B, Parainfluenza Virus 1, Parainfluenza Virus 2, Parainfluenza Virus 3, Parainfluenza Virus 4, Rhinovirus/Enterovirus, Respiratory Syncytial Virus A+B, *Bordetella pertussis, Chlamydophila pneumoniae and Mycoplasma pneumoniae*) (supplement 2).

Total RNA was extracted using the QIAamp Viral RNA Mini Kit (QIAGEN, Germany), and sent for the metagenomic sequencing. During metagenomic sequencing, RNA was reverse-transcribed into cDNA, with which sequencing library was prepared. The qualified library was sequenced on an Illumina Nextseq using a single-end mode (1x75bp). After low-quality reads and reads derived from human genome sequences were removed, *de novo* assembly was done using SPAdes(v3.1). Sequenced reads were aligned to a curated database for taxonomic classification, SARS-COV-2 and other pathogen identification as described previously^16^.

### Statistical Analysis

Categorical variables were described as frequency rates, and continuous variables were described using median, and interquartile range (IQR) values. Means for continuous variables were compared using independent group t tests when the data were normally distributed; otherwise, the Mann-Whitney test was used. Proportions for categorical variables were compared using the χ2 test, although the Fisher exact test was used when the data were limited. All statistical analyses were performed using SPSS (Statistical Package for the Social Sciences) version 13.0 software (SPSS Inc). For unadjusted comparisons, a 2-sided α of less than 0.05 was considered statistically significant.

### Phylogenetic analysis

Thirty genomes of coronal viruses were downloaded from NCBI and 6 Wuhan SARS-COV-2 genomes released by China through GISAID (https://www.gisaid.org/)“www.gisaid.org/) for phylogenetic analysis. Phylogenetic trees were constructed based on the N gene sequences by means of the maximum-likelihood method^17^. A total of 10 samples which had sequence coverage of over 80% were included in the analysis. Alignment of multiple sequences was performed with the ClustalW program (MEGA software, version 7.0.14)^18^.

## Results

Our study enrolled 53 patients with suspected NCP. Twenty patients were finally laboratory-confirmed with NCP, among whom 14 patients were positive for the first admission SARS-COV-2 RT-PCR, 3 patients turned to positive in the second test after the first negative result. Three other patients were diagnosed through mNGS while both RT-PCR reported negative (Figure 1, Table 1, Supplement 1). The virus load of the 17 NCP patients varied from 2.0 to 2.1×10^6^ copies/μL (Supplement 2).

**Table 1.** Diagnostic efficacy of SARS-CoV-2 RT-PCR, mNGS, direct antigen Flu A+B test,multiplex PCR for other respiratory infection pathogens.

|  | Diagnostic methods | Laboratory-confirmed NCP (n=20) | Laboratory-confirmed non NCP (n=33) |
| --- | --- | --- | --- |
| **2019-nCoV identification** | Suspected chest CT signs | 20/20 | 20/33 |
|  | First time SARS-CoV-2 PCR positive | 14/20 | 0/33 |
|  | Second time SARS-CoV-2 PCR positive | 3/6 | 0/33 |
|  | mNGS positive for SARS-CoV-2 | 20/20 | 0/33 |
| **Other respiratory infection pathogens** | Direct antigen Flu A+B test positive | 0/20 | 0/33 |
|  | Multiplex PCR positive for other pathogens | 5/20 | 7/20 |
|  | mNGS positive for other pathogens | 11/20 | 23/33 |

**Figure 1.**
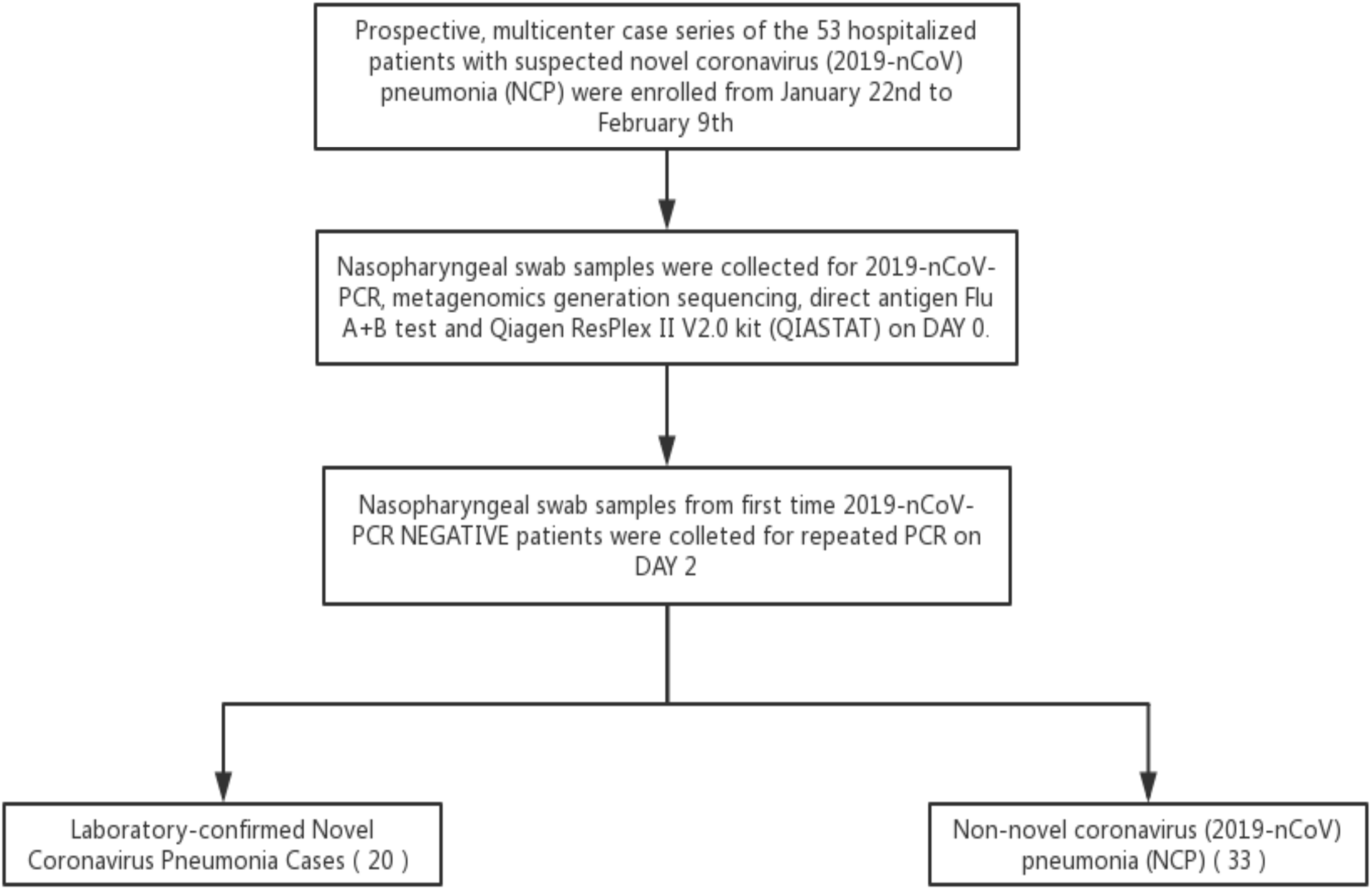
Patient recruitment flowchart.

Among the conformed cases, 16 NCP patients had a travel history from Hubei Province and 3 patients had close contact with patients with fever or respiratory symptoms from Hubei Province or confirmed cases with 14 days on diseases onset. One patient didn’t have any typical epidemiology history. Half of the NCP patients were male, with a median age of 37 years. The most common symptoms at onset of NCP were fever (16 [80%]), dry cough (11 [55%]), diarrhea (3 [15%]), and headache (3 [15%]). Less common symptoms were fatigue, abdominal pain, and vomiting. Three patients were diagnosed with severe COVID-19 pneumonia. Importantly, the symptoms of NCP patients were very similar with non-NCP patients in our study. The blood counts of enrolled NCP patients showed less white blood cells, neutrophils and platelet count than non-NCP patients (P<0.05). Levels of aspartate amino transferase were increased in 3 (15%) of 20 NCP patients. (Supplement 1).

Abnormalities in chest CT images were detected among all patients. According to CT scan, 12 (60%) NCP patients showed bilateral pneumonia with just 8 (40%) patients showing unilateral pneumonia. Most non-NCP patients showed unilateral or bilateral patchy shadowing. TFour non-NCP and 2 NCP patients showed ground-glass opacity, respectively. CT image of other patients were subsegmental areas of consolidation. In our study, chest CT images of early stage NCP patients showed unilateral or bilateral ground glass opacity, which were similar to some non-NCP images of patients with RSV, mycoplasma and parainfluenza virus. The findings of progressive stage chest CT images were bilateral multiple lobular and subsegmental areas of consolidation (Figure 2).

**Figure 2.**
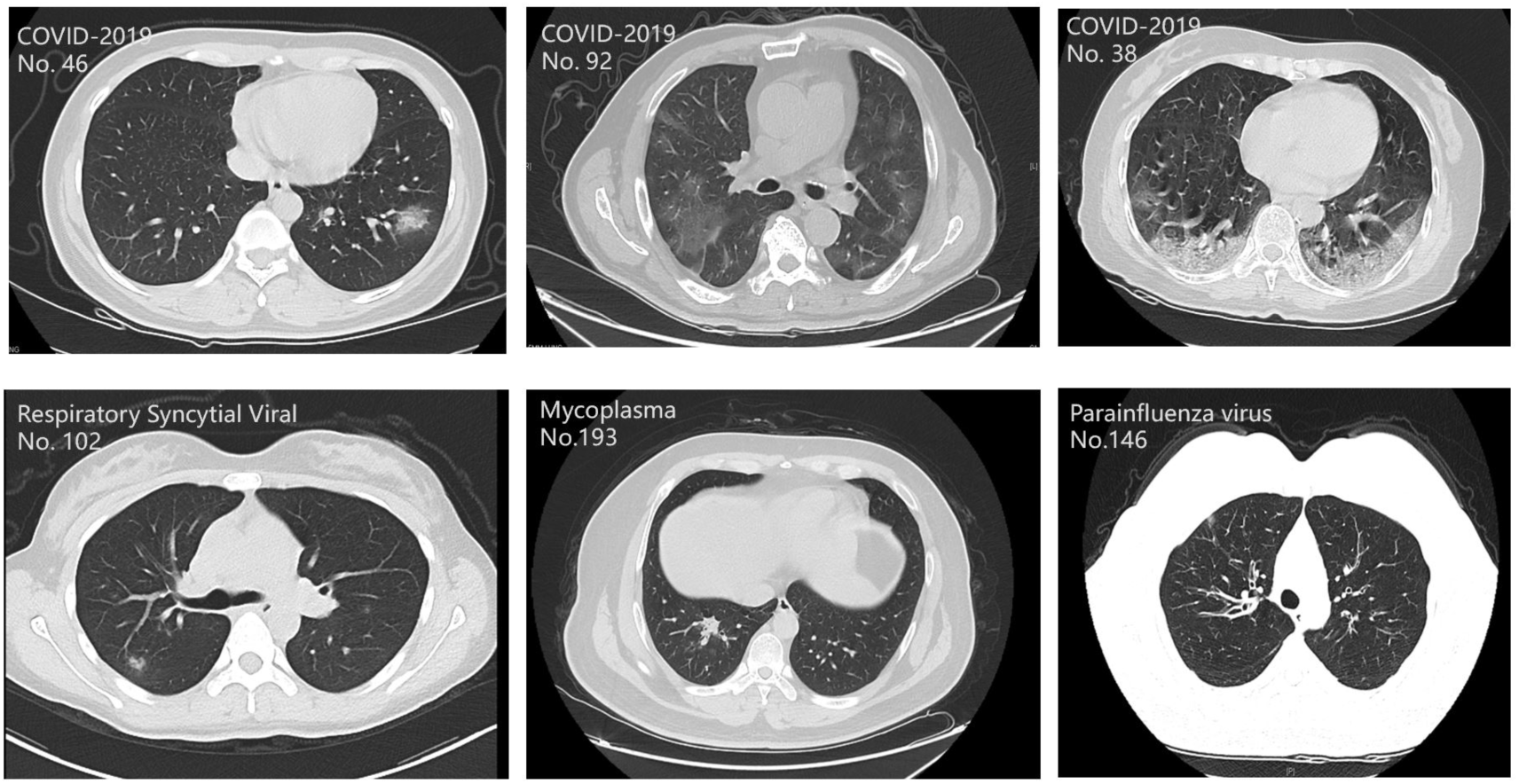
Chest computed tomographic Images of patients with SARS-CoV-2 and other pathogens.

According to the QIASTAT and metagenomic sequencing results, we found 11 co-infection cases among 20 NCP cases, including viruses such as influenza A/B, rhino/enterovirus, respiratory syncytial viral and etc., and other atypical pathogen, fungi and bacteria synchronously (Figure 3). Among the 33 laboratory-confirmed non-NCP patients, QIASTAT and metagenomic sequencing reported positive in 7 and 23 patients, respectively. Pathogen distribution of non-NCP cases was shown in Figure 3.

**Figure 3.**
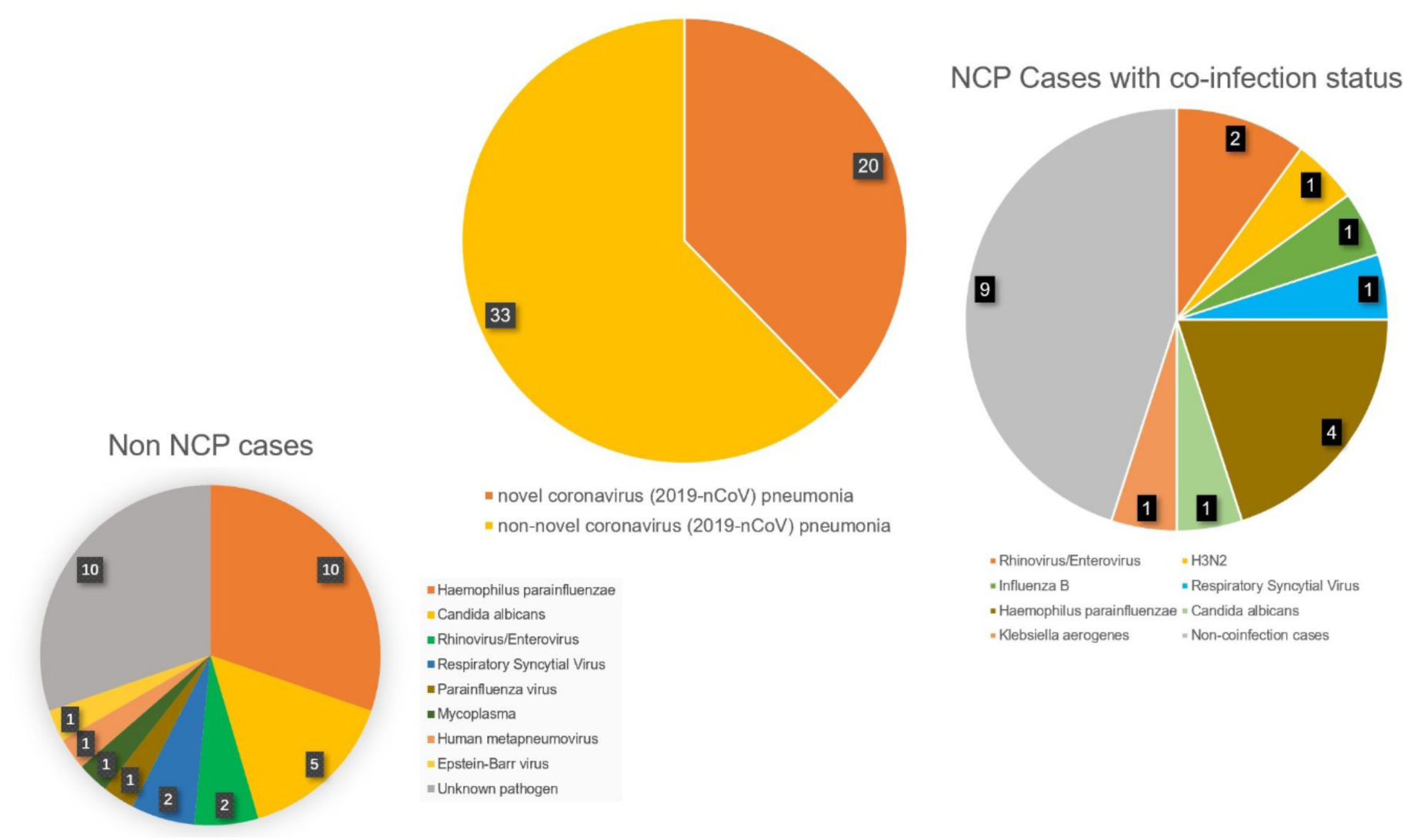
Pathogen distribution of 53 patients with suspected NCP.

Therefore, in the diagnosis of unexplained pneumonia, positive respiratory pathogen results cannot serve as an evidence for exclusion of SARS-COV-2 infection. Currently, non-SARS-COV-2 pathogens are still very common in suspected patients in Eastern China region.

We applied metagenomic sequencing for SARS-COV-2 identification in these samples. Out of the 53 suspected samples, we were able to identify 20 SARS-COV-2-positive cases, all 17 of RT-PCR positive samples were mNGS positive and 3 repeated RT-PCR negative patients were also tested positive during mNGS. The metagenomics results showed median reads of all SARS-COV-2-positive cases per million of 99 (1-688,541), a median genome coverage of 35.0% (1.2-99.9%) and a median sequencing depth of 24x (1x-7453) (Supplement 3). The SARS-COV-2 reads showed by metagenomics are positively correlated with SARS-COV-2 copies by RT-PCR (Spearman correlation, r=-0.736, *P*<0.001). The great degree of variations shown in these sequencing outputs suggests a broad range of viral titer in SARS-COV-2 infections.

Phylogenetic analysis was performed to investigate the genetic relationship between SARS-COV-2 and other pathogenic coronavirus, as well as the diversity among our SARS-COV-2 strains. Consistent with previous reports, SARS-COV-2 was identified as a new species that’s most similar to SARS-CoV. The SARS-COV-2 strains sequenced in our cases were highly conserved with the original Wuhan strain. Genetic variations were further analyzed in 10 cases with sufficient coverage and depth of the N gene. Among which, only one single nucleotide mutation (g605a) was identified in two of the samples which resulted in a non-synonymous S203N change in the N gene, suggesting a relatively low level of genetic polymorphism in our SARS-COV-2 cases (Figure 4).

**Figure 4.**
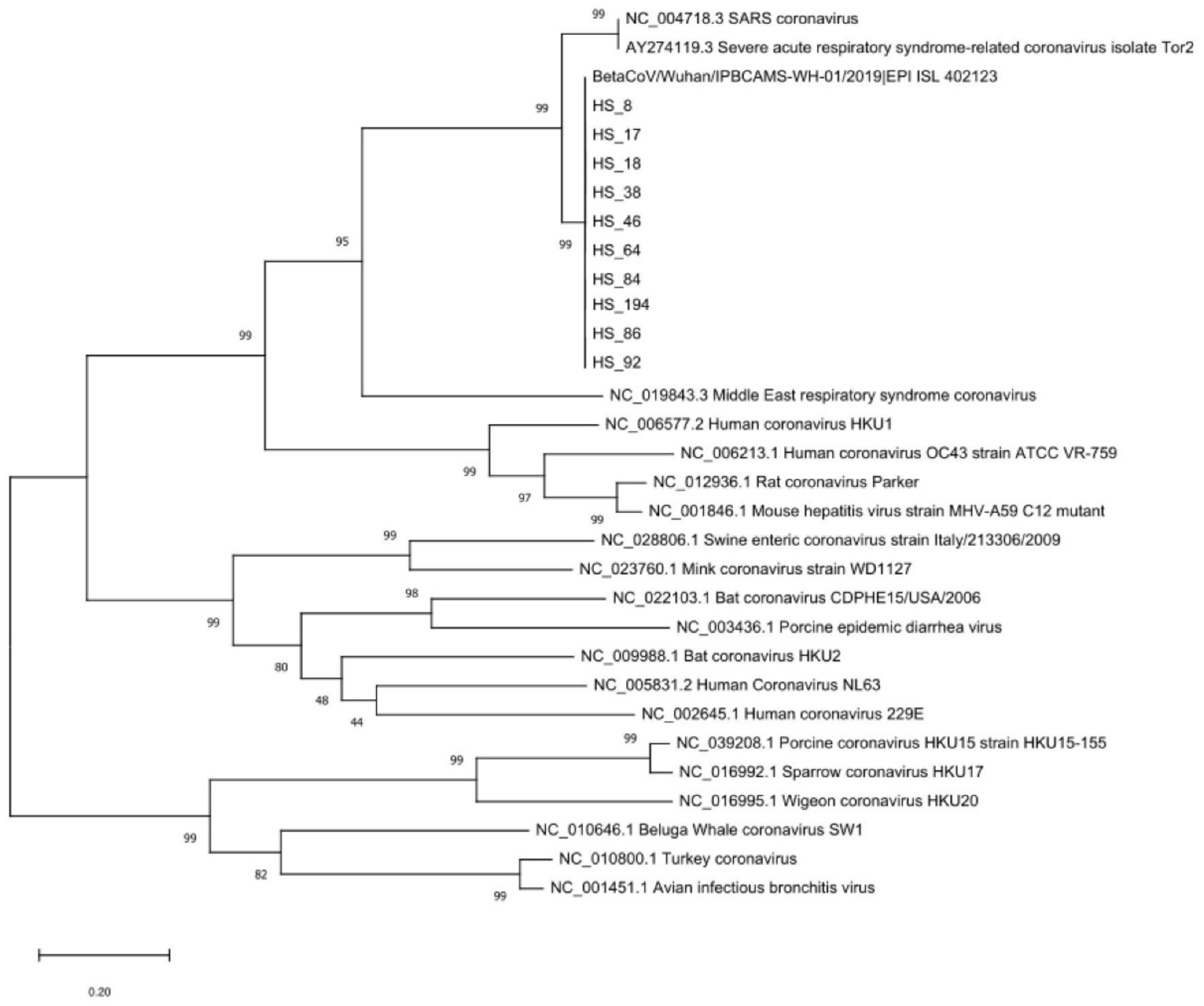
Phylogenetic analysis of SARS-CoV-2 positive samples based on the N gene sequences.

## Discussion

In our study, we enrolled 53 suspected cases and 20 cases were finally laboratory-confirmed with NCP, among which 11 patients were also co-infected with other pathogens. Other causative pathogens were detected through multiplex respiratory PCR panel and mNGS, and in the non-NCP groups, some pathogens, such as RSV, parainfluenza virus may share very similar radiological patterns as the NCP group, including bilateral multiple lobular, ground-glass opacity, and consolidation. This indicated that for regions in Eastern China, pathogens other than SARS-COV-2 still composed of approximately 60% of the suspected unexplained pneumonia; therefore, chest CT alone could not be used as a diagnostic criterion for SARS-COV-2 infections. A combination of chest CT and other diagnostic methods is essential to decrease the chance of misdiagnosis in areas outside Hubei Province.

In the previous articles, researchers found severe ICU patients had coinfections of bacteria and fungi^19^. It probably occurred secondary to severe ARDS or large dose or long time of glucocorticoid therapy. The common pathogen included *A. baumannii, K. pneumoniae, A flavus, C. glabrata*, and *C. albicans*. However in our study, we found 5 co-infection virus cases other than the common bacterial and fungi, including influenza, rhino/enterovirus, respiratory syncytial viral and etc, suggesting that virus co-infection may be the common phenomena in winter pneumonia cases. Thus, positive pathogen results (especially virus and atypical pathogens, which is a type of common pathogens detection in unexplained pneumonia) cannot be the exclusive criteria to COVID-2019 during unexplained pneumonia diagnosis.

In the laboratory-confirmed NCP cases, 15% of the nasopharyngeal swab specimens were negative for RT-PCR on the first time and turned to positive on the second time, while another 15% of the specimens tested negative both time and was finally diagnosed through metagenomics sequencing. The result suggested that the current RT-PCR for SARS-COV-2 indeed had at least 15% of false negative rate. However, these 3 mNGS positive but RT-PCR negative samples had a relatively low sequences reads, which suggested low viral loads in the upper respiratory tract and the possibility low contagious risk for these patients.

The reasons lies behind this phenomenon may be due to several causes. One of the influencing factors is the sampling techniques. We found very high copies of NCP virus in the deep nasopharyngeal swab sample of the patient No. 18, whose first nasopharyngeal swab collected one day before by another physician was negative, which suggests that standard sampling strictly according to the protocol may increase the sensitivity of the specimens. Also, most of the cases published until now tends to use nasopharyngeal swab specimens for nucleic acid tests^19,20^. Our study revealed that for some NCP patients with very mild pneumonia, their viral loads and mNGS sequences reads from the nasopharyngeal swab specimen could be as high as 2×10^6^ copies/ul and 21,938,215 (patient No.8, seen in Supplement 1), the highest among all patients. Meanwhile, some patients having severe symptoms had relatively low virus loads detected form nasal swab samples. For example, patient no.92 were admitted into hospital with severe pneumonia and an oxygen saturation of below 93%; however, the SARS-COV-2 viral loads of his sample is 3.1X10^3^ copies/μl while mNGS detected 1,216 unique reads, relatively lower than other NCP patients (Supplement 1). A possible explanation could be that NCP patients’ lesions tends to be located in the lower respiratory tract, causing the RT-PCR positivity of the upper respiratory tract sampling to be reduced. Other factors such as RNA extraction efficacy, quality of kit or potential interference of multiple primers could also affect the detection of RT-PCR assay. Possible mutation in primer or probes regions could lead to false-negative detection, albeit these regions are very reserved. Although from WGS data, we haven’t found any mutation in the primer regions in these patients, however, such risk always exits. The primer design also partially relies on the chosen gene. *Orf1ab* and S gene are very specific genes that can effectively distinguish COVID-19 from other coronaviruses (including SARS and SARS like viruses), which have been used to design specific PCR detection primers. N and E genes of the SARS-CoV-2 may form cross-reaction with other coronaviruses.

Metagenomic sequencing was the method by which SARS-COV-2 was initially identified. Despite its longer turnaround time and higher cost, mNGS demonstrated a satisfying level of sensitivity when compared to PCR assays in our study. Moreover, the nature of unbiased sequencing offers higher tolerance for genetic drift in the target pathogens, whereas certain changes in sequence can result in critical impact on the assay efficiency^21,22^. Along with targeted assays, mNGS should serve as an indispensable tool for sensitive and close monitoring of the molecular evolution for SARS-COV-2. Such genetic information will offer valuable insights not only for public health management, but also for contentious optimization of diagnostic assays^23,24^.

Therefore, as the SARS-COV-2 epidemic is currently entering into a critical stage, both within China and over the global^25^, according to the results of our study, the following improvements in molecular diagnosis are recommended in the optimized diagnostic flow diagram [Figure 5]. First of all, repeated sampling could increase the positivity, and especially during clinical situations where epidemiological history and clinical manifestations highly indicate NCP, a third or fourth sampling may be helpful when resources are available. Second, our study showed that mNGS showed reliable and stable detection ability in the NCP diagnosis, and may facilitate to solve diagnostic difficulties during important or critical clinical cases and low positive RT-PCR results. What’s more, in order to circumvent possible mutations in RT-PCR primers region, more than 1 gene target was recommended to amplify when designing RT-PCR assays [25]. Whole Genome Sequencing (WGS) method can overcome the mutation problems which cause false-negative result in RT-PCR. Therefore, we used this method in all enrolled patients and should be continue using for mutation surveillance. Lastly, lower respiratory tract specimens might provide higher positivity, due to that higher viral loads may be detected there. Therefore, physicians might collect such samples through bronchoscopy or trachea cannula when under through medical protection.

**Figure 5.**
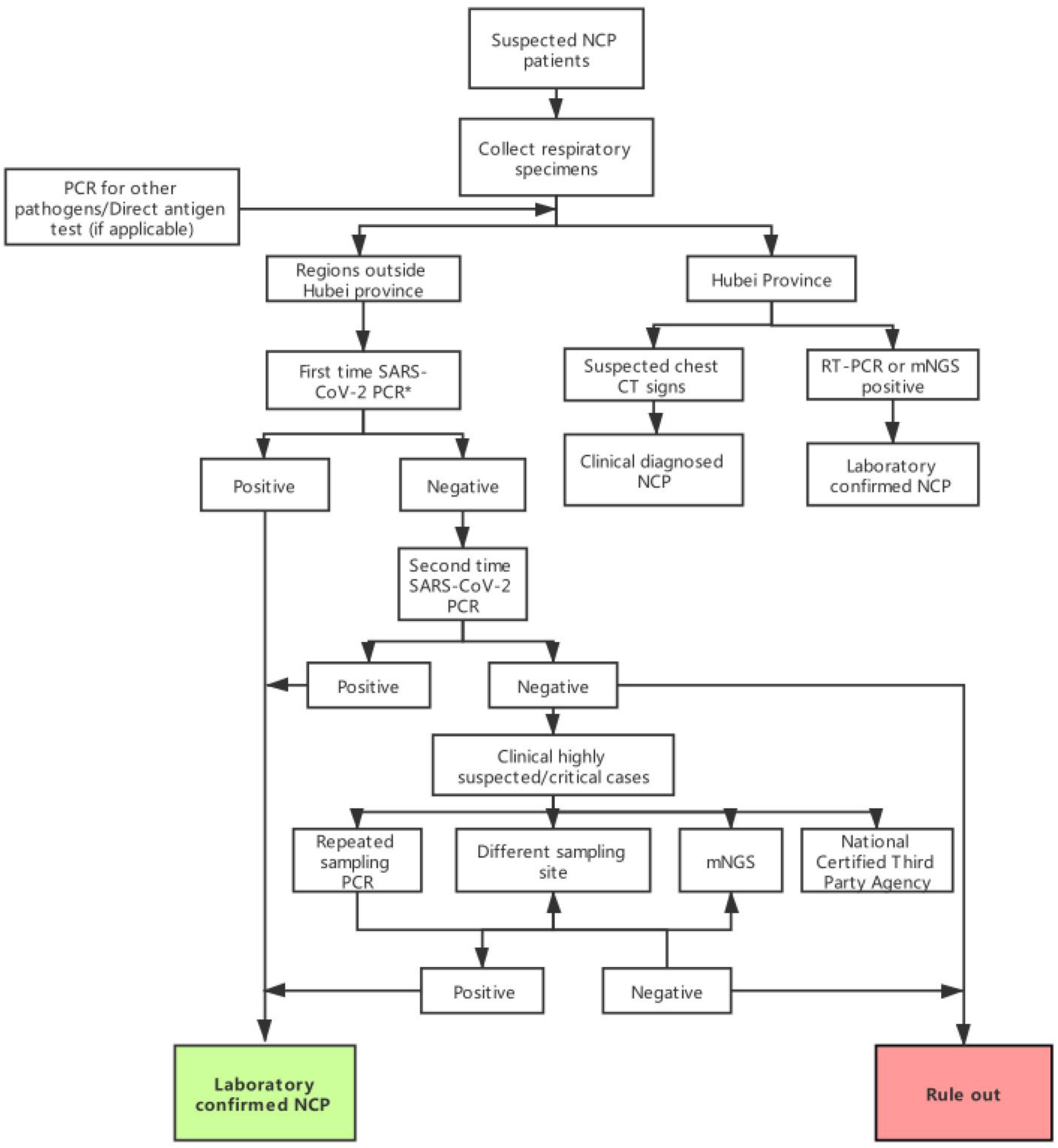
Recommended optimized diagnostic flow diagram. * Should other tests such as ELISA, CRISPR, LAMP, etc. be validated for SARS-CoV-2 detection, these tests may also be used in the first-step diagnosis.

In the future, rapid serological tests for antigen and antibodies, targeted molecular assays (e.g. PCR- or CRISPR-based ones^26^) are currently under development, and if successfully validated, these methods could further be used in combination with other diagnostic method to assist in clinical practice.

The study has a limitation of a relatively small sample size. More studies are needed to further validate our findings. Also, sputum, lower respiratory tract, serum and fecal samples were not collected for our laboratory-confirmed patients in our study, and collection of the above samples for a larger cohort would help to improve our diagnostic strategy.

Our study discovered that, in low epidemic regions outside Hubei province such as Eastern-China region, the current reliable first approach to the unexplained pneumonia should be a combination of chest CT, SARS-COV-2 RT-PCR and multi-plex PCR (rapid influenza antigen tests and RT-PCR could also be used as a substitute). Chest CT alone could not precisely diagnose COVID-2019 due to similar radiological presentations. Co-infection of other pathogens with SARS-COV-2 exists and should be acknowledged. What’s more, different sampling approaches may also affect the results, therefore, in highly suspected cases but with two negative SARS-COV-2 RT-PCR results, repeated sampling or change of sample type could increase the positive rate. Metagenomics sequencing could further facilitate during critical clinical cases and monitor the molecular evolution for SARS-COV-2.

## Data Availability

All data referred in the manuscript is available in the paper.

## Acknowledgement

This study is sponsored by the National Science and Technology Major Project of China (2018ZX10305-409-001-003). We acknowledge all health-care workers involved in the diagnosis and treatment of patients in Eastern China; we thank the Shanghai Municipal Center for Disease Control and Prevention, and Jian District Center for Disease Control and Prevention; We acknowledged BGI Genomics for their assistance in the analysis of molecular diagnosis.; we thank anonymous Shanghai citizens and companies for their donation of personal protective equipment.

---

**Supplement 1.** Clinical characteristics of 53 patients with suspected SARS-CoV-2 pneumonia.

|  | NCP positive cases (N=20) | NCP negative cases (N=33) | P |
| --- | --- | --- | --- |
| **Age, median (IQR)** | 37(33.75-50.5) | 39 (30.5-43) | 0.882 |
| **Sex** |  |  |  |
| Female | 10 (50.0%) | 17 (51.5%) | 0.915 |
| Male | 10 (50.0%) | 16 (48.5%) |  |
| **Signs and symptoms** |  |  |  |
| Fever | 16 (80.0%) | 17 (51.5%) |  |
| Dry cough | 11 (55.0%) | 19 (57.6%) |  |
| Diarrhea | 3 (15.0%) | 4 (12.1%) |  |
| Fatigue | 2 (10.0%) | 2 (6.1%) |  |
| Headache | 3 (15.0%) | 1 (3.0%) |  |
| Vomiting | 1 (5.0%) | 0 (0.0%) |  |
| Abdominal pain | 1 (5%) | 0 (0.0%) |  |
| **Laboratory Findings, median (IQR)** |  |  |  |
| White blood cell count, $\times 10^9/L$ | 4.21 (4.03-6.70) | 8.38 (6.27-9.64) | 0.001 |
| Neutrophil count, $\times 10^9/L$ | 2.88 (2.25-4.63) | 4.94 (4.10-6.61) | 0.000 |
| Lymphocyte count, $\times 10^9/L$ | 1.21 (0.91-1.46) | 1.54 (1.09-2.12) | 0.077 |
| Haemoglobin, g/L | 145 (131.25-158) | 143 (128-155) | 0.528 |
| Platelet count, $\times 10^9/L$ | 199(146-248.25) | 245(197-279) | 0.017 |
| C-reactive protein, mg/L | 9.68(1.63-25.83) | 5.84(2.93-16.74) | 0.340 |
| Alanine aminotransferase, U/L | 21.50 (15-31.25) | 46 (36-60) | 0.000 |
| Aspartate aminotransferase, U/L | 23 (18.50-27) | 26 (23-34) | 0.006 |
| Total bilirubin, mmol/L | 6.70(6.15-7.75) | 7.8(7.08-10.35) | 0.153 |
| Albumin, g/L | 43(40.50-46.95) | 46(44.23-50) | 0.083 |

